# ECG spectrogram-based deep learning model to predict deterioration of patients with early sepsis at the emergency department: a study from the Acutelines data- and biobank

**DOI:** 10.64898/2026.03.26.26349371

**Authors:** Raymond J. van Wijk, Anna D. Schoonhoven, Leanne de Vree, Sanne Ter Horst, Chetan Gaidhane, Juan Miguel Lopez Alcaraz, Nils Strodthoff, Jan C. ter Maaten, Hjalmar R. Bouma, Jie Li

**Author notes:** Corresponding author: R.J. van Wijk.

## Abstract

**Purpose:** Early recognition of deterioration in patients with suspected infection at the emergency department (ED) is important. Current clinical scoring systems show limited discriminative performance for early deterioration. Continuous electrocardiogram (ECG) recordings may offer additional dynamic physiological information that can enhance early prediction of deterioration in patients with suspected infection.

**Methods:** We developed a multimodal, ECG-derived spectrogram-based pipeline to predict deterioration within 48 hours of ED admission. We used the first 20 minutes of ECG recordings for the spectrograms. We compared the model with the National Early Warning Score (NEWS), quick Sequential Organ Failure Assessment (qSOFA), a baseline model with vital parameters, sex, and age, and a Heart Rate Variability (HRV) derived model.

**Results:** In this study, 1321 patients were included, of whom 159 (12%) deteriorated. The multimodal model combining baseline data with spectrograms showed the best overall performance, with an Area Under the Receiver Operating Characteristic (AUROC) of 0.788, followed by the baseline model (age, sex, triage vitals) alone, with an AUROC of 0.730. The HRV-only model and the qSOFA showed the lowest performance (AUROC 0.585 and 0.693, respectively).

**Conclusion:** This study shows that ECG-derived multimodal spectrogram models outperform those based solely on vital signs and HRV features, as well as established clinical scores such as NEWS and qSOFA. Spectrogram analysis represents a promising approach to enhance early risk stratification and support clinical decision-making for patients with suspicion of infection in the ED.

## Introduction

Sepsis is a life-threatening condition resulting from a dysregulated host response to infection and is responsible for 20 million deaths worldwide each year [1–4]. Early recognition is important to initiate timely and appropriate care [5]. However, identifying future deterioration of patients presenting at the Emergency Department (ED) with infection of early sepsis remains challenging, as early clinical manifestations are often non-specific and highly variable between individuals [6,7]. This clinical heterogeneity limits clinicians’ ability to reliably distinguish patients at risk of future clinical deterioration. Consequently, delays in treatment initiation and decision regarding escalation of care are common. Among patients admitted with suspected infection, approximately 20-30% deteriorate within 48 hours after hospitalization [8,9]. This poses a challenge for physicians in the ED, who must distinguish between deteriorating and non-deteriorating patients. Health care professionals in the ED commonly rely on early warning scores, such as the National Early Warning Score (NEWS) and NEWS2, and sepsis-specific tools, such as the quick Sequential Organ Failure Assessment (qSOFA), to identify patients at risk of clinical deterioration [4,10,11]. These tools are designed to support early recognition and guide treatment and disposition decisions. However, the prognostic accuracy remains limited, as they are primarily based on discrete measurements of vital signs obtained at fixed time points [12,13]. Although repeated assessments and temporal changes in NEWS, qSOFA and individual vital signs have been shown to moderately predict subsequent deterioration [9,14], these approaches still depend on intermittent data collection and manual interpretation. As a result, important short-term physiological fluctuations, including autonomic and cardiovascular dynamics, may go undetected, limiting their usefulness for timely, individualized risk assessment.

In addition to intermittent measurements, continuous physiological electrocardiogram (ECG) monitoring provides assessment of beat-to-beat cardiovascular dynamics. In particular, Heart Rate Variability (HRV), which is derived from the ECG beat-to-beat interval, reflects autonomic regulation and captures complex temporal patterns in cardiovascular function [15,16]. Previous studies have shown that HRV characteristics differ between patients who deteriorate and those who remain clinically stable [17]. For instance, low-frequency components in HRV are related to baroreflex sensitivity and blood pressure control, both known to be affected in (early) sepsis due to vasodilation and vascular leakage [18,19]. These findings suggest that subtle changes in ECG-waveforms may precede clinical deterioration. Furthermore, model performance for predicting deterioration improves when waveforms are added to clinical data [20]. However, HRV analysis is typically performed using predefined time windows, most commonly five-minute intervals, which limits temporal resolution [21]. Moreover, HRV-based approaches rely on extensive pre-processing and predefined feature extraction [16,21], which constrain their applicability for real-time clinical decision-making. In summary, HRV analysis at the ED is limited by (i) temporal resolution, (ii) reliance on predefined features, and (iii) feasibility for real-time ED use.

ECG spectrograms provide a way to represent dynamic waveform characteristics with high temporal resolution. This technique provides a visual representation of ECG signals that combines time and frequency components into a single 2D image, reflecting how spectral components evolve over time [22]. Hypothetically, spectrograms could capture long-range dependencies more effectively than raw waveforms. Spectrogram-based analysis of ECG-signals has shown value in other fields, such as arrhythmia detection [23,24]. However, interpretation of spectrograms remains challenging, particularly in the absence of clearly defined visual markers. Modern deep learning approaches automatically extract and learn hidden features, eliminating the need for manual processing, feature extraction, and interpretation [23,24]. Applied to patients with suspected infection or sepsis in the ED, such methods facilitate early identification, by capturing non-stationary dynamics, of individuals at risk of clinical deterioration. It is unknown how such spectrogram-based deep learning approaches compare with conventional methods such as basic vital sign measurements, NEWS and qSOFA, and ECG-based HRV analysis. In this study, we explore an unbiased approach to predict clinical outcomes for patients with suspected infection in the ED by applying deep learning to ECG-derived spectrograms of the first 20 minutes after arrival. The aim was to develop a proof-of-concept framework for applying deep-learning-based spectrogram analysis in the ED, evaluate its performance relative to conventional scoring systems and HRV, and lay the foundation for future multimodal deterioration prediction models. We hypothesize that this spectrogram-based deep learning approach has better predictive performance for subsequent clinical deterioration of patients arriving at the ED than NEWS, qSOFA, and traditional HRV analysis.

## Methods

### Study design

We conducted a post-hoc analysis of prospectively obtained data by the Acutelines data- and biobank at the Emergency Department of the University Medical Center Groningen (UMCG; Groningen, the Netherlands). Acutelines is a multi-disciplinary prospective hospital-based cohort study that includes patients 24/7 and examines the complete acute patient journey of patients admitted to the ED of the University Medical Centre Groningen (UMCG), a tertiary care teaching hospital in the Netherlands. Detailed information about the cohort and participant selection can be found elsewhere [25,26]. Participants were asked for written informed consent, when applicable by proxy. The Acutelines cohort study is approved by the medical ethics committee of the UMCG, the Netherlands, and registered under trial registration number NCT04615065 at ClinicalTrials.gov [26]. This study was approved by the institutional review board of the UMCG (#18939).

### Study population

Adult patients (≥18 years) were included if admitted through the emergency department (for internal medicine, rheumatology, gastroenterology, pulmonology, or emergency medicine) with a clinical suspicion of infection as determined by routine ED practice. Suspicion could arise from clinical judgement, abnormal vital signs (e.g., ≥2 SIRS or ≥2 qSOFA criteria), or infection-related triggers such as blood cultures drawn or abnormal temperature, particularly in high-acuity patients.

The exclusion criteria were known pregnancy (i), patient admitted to another hospital (ii), absent or low-quality ECG-data (iii), and any repeated visit of the same patient (iv). For this study, we used the first 20 minutes of the ECG waveform recorded after arrival at the ED, which aligns with the moment of triage when the patient’s clinical condition is typically assessed based on vital parameters, before laboratory parameters are available. This allows fair comparison to current triage practices, while maintaining a timely intervention window (e.g. early antibiotics). Patients were excluded if no single heart rate, oxygen saturation, or blood pressure measurements were taken at the ED.

### Endpoints

The primary endpoint was clinical patient deterioration, defined as the composite outcome of ICU admission within 48 hours and/or in-hospital mortality within 48 hours after ED admission. This time window was selected because infection-related clinical deterioration typically occurs within the first 48 hours following ED admission [8].

### Data acquisition and preprocessing

Data were retrieved from patients’ electronic health records, and ECG signals were continuously recorded at the ED. Baseline model data consisted of age, sex, heart rate (HR), respiratory rate (RR), peripheral oxygen saturation (SpO2), systolic blood pressure (SBP), diastolic blood pressure (DBP), temperature (temp), and Glasgow Coma Scale (GCS). Missing Glasgow coma scores were imputed as 15, respiratory rates as 12 breaths per minute, and temperatures as 37 °C. These imputation values reflect normal physiological ranges and were chosen because undocumented measurements in electronic health records are generally assumed to represent normal findings.

NEWS and qSOFA were calculated from triage vital parameters using the Acutelines datatoolbox version 1.1.0 [27]. A cutoff of 5 was used for NEWS, and a cutoff of 2 was used for qSOFA predictions in comparisons to the developed models. Raw 500 Hz ECG waveforms were acquired from the Philips IntelliVue (Philips, Amsterdam, the Netherlands) bedside patient monitor and automatically stored at UMCG’s proprietary waveform platform. The first 20 minutes of waveform recordings were selected and extracted in high resolution for analysis and used for generating spectrograms and extracting HRV features.

### Statistical analysis and model development

Baseline vital parameters and demographics were tested for normality using the Shapiro-Wilk test. For normally distributed data, mean and standard deviation (SD) were calculated. For non-normally distributed data, the median and interquartile ranges (IQR) were calculated. Differences between the two groups were tested with the Mann-Whitney U test. A p-value < 0.05 was considered significant. Demographic statistical analyses were performed using R version 4.2.2 and model development was done in Python 3.13.3 using libraries scikit-learn version 1.6.1 and PyTorch version 2.7.1. All Receiver Operating Characteristic (ROC) figures were generated with Matplotlib version 3.10.1.

We developed a modular multimodal deep learning framework to integrate heterogeneous physiological information derived from ECG signals, routine vital signs, and baseline demographics. The framework consists of modality-specific encoders followed by a feature-level fusion module, enabling direct comparison between single-modality and multi-modal models under the unified training and evaluation protocol. Three model variants were investigated:

i. a spectrogram-only model, Convolutional Neural Network (CNN)-based
ii. a baseline (age, sex, triage vitals) or HRV feature-only model, Multilayer Perceptron (MLP)-based, and
iii. a multi-modal model combining baseline with HRV features or spectrograms.

An overview of the data pipeline and general model architecture is shown in *Figure 1*, and the complete model architecture is available in *Supplemental Figure A1*. Detailed methods on the model development are available in the *Supplemental Section A*. We additionally investigated tree-based models for tabular data, as shown in Supplemental Section B. Performance metrics Area Under the Receiver Operating Characteristic (AUROC), Specificity, Positive Predictive Value (PPV), Negative Predictive Value (NPV), F1-score, F2-score were reported at a fixed sensitivity of 80% to allow fair comparison, including 95% confidence intervals (CI).

**Figure 1:**
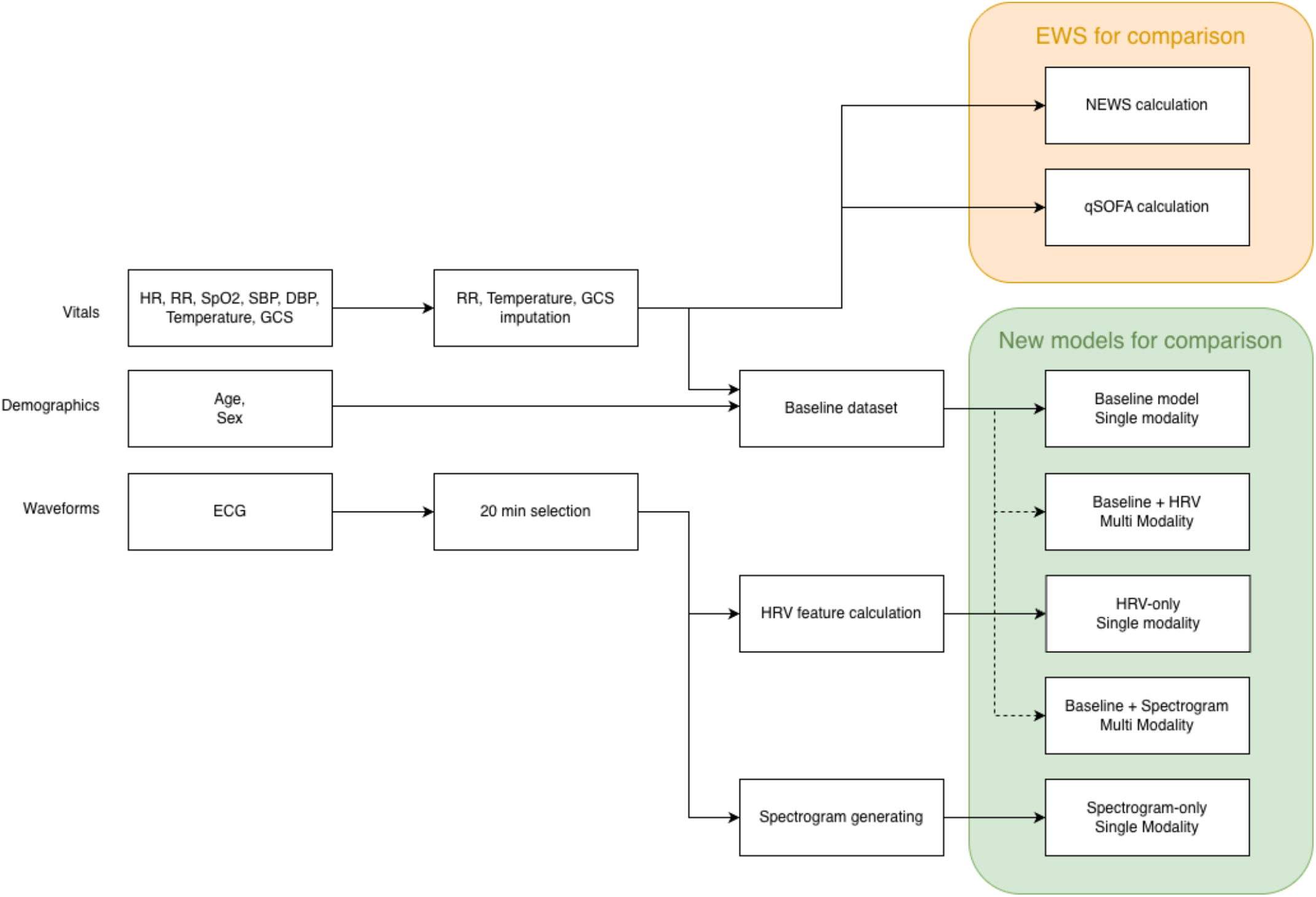
Overview of the data pipeline and general model architecture.

## Results

### Study population

Of the 1321 included patients, 159 (12%) deteriorated within 48 hours of ED admission, defined as ICU admission or death, while 1162 (88%) did not. Baseline and outcome study population characteristics are shown in *Table 1*. Deteriorated patients were on average younger, with a median age of 61 years versus 67 years in the non-deteriorated group (p<0.05). Overall, deteriorated patients received a higher triage urgency (Emergency Severity Index) upon arrival at the ED and were more often transported by ambulance. Vital parameters at triage differed between the groups on heart rate (106 bpm in deteriorated patients vs 98 bpm in non-deteriorated patients), respiratory rate (23 /min vs 20 /min), SBP (116 mmHg vs 130 mmHg) as well as mean arterial pressure (87 mmHg vs 95 mmHg) and SpO2 (95% vs 96%). Although specific comorbidities did not differ between the two groups, the Charlson Comorbidity Index (CCI) was lower among deteriorated patients (3 vs 4). Several laboratory values, including creatinine (105 µmol/L vs 87 µmol/L), ASAT (41 U/L vs 28 U/L), ALAT (29 U/L vs 23 U/L), PaO2 (11.2 kPa vs 8.9 kPa), and lactate (2.5 mmol/L vs 1.4 mmol/L), were higher upon ED arrival in the deteriorated group as compared to non-deteriorated patients.

**Table 1:** Overview of baseline and outcome characteristics of the population, stratified by outcome: not deteriorated and deteriorated. Deterioration was defined as ICU admission and/or death within 48 hours. Results are reported as n (%) for categorical variables and median [IQR] for continuous variables. Reported p-values are for differences tested with the Mann-Whitney U test.

|  | Not deteriorated | Deteriorated | p |
| --- | --- | --- | --- |
| n | 1,162 | 159 |  |
| Age (years) | 67 [55, 75] | 61 [51, 71] | <0.05 |
| Sex |  |  | 0.516 |
| male | 688 (59.2) | 99 (62.3) |  |
| female | 474 (40.8) | 60 (37.7) |  |
| Transport method |  |  | <0.05 |
| Own transport | 329 (28.3) | 15 (9.4) |  |
| Ambulance | 825 (71.1) | 142 (89.3) |  |
| HEMS | 3 (0.3) | 2 (1.3) |  |
| In-hospital emergency | 4 (0.3) | 0 (0.0) |  |
| Emergency Severity Index (%) |  |  | <0.05 |
| 1 (Red) | 11 (1.0) | 35 (22.9) |  |
| 2 (Orange) | 383 (33.5) | 82 (53.6) |  |
| 3 (Yellow) | 749 (65.5) | 36 (23.5) |  |
| **Vital parameters at triage** |  |  |  |
| Temperature (°C) | 37.1 [36.5, 38.0] | 36.5 [36.0, 37.4] | <0.05 |
| Heart rate (bpm) | 98 [83, 113] | 106 [90, 122] | <0.05 |
| Respiratory rate (bpm) | 20 [17, 25] | 23 [19, 30] | <0.05 |
| SBP (mmHg) | 130 [114, 148] | 116 [93, 140] | <0.05 |
| DBP (mmHg) | 77 [66, 89] | 74 [57, 90] | <0.05 |
| MAP (mmHg) | 95 [84, 107] | 87 [71, 103] | <0.05 |
| SpO<sub>2</sub> (%) | 96 [94, 98] | 95 [92, 98] | <0.05 |
| GCS | 15 [15, 15] | 15 [13, 15] | <0.05 |
| **Comorbidities** |  |  |  |
| Charlson Comorbidity Index | 4 [2, 6] | 3 [1, 5] | <0.05 |
| **Infection etiology and source** |  |  |  |
| No confirmation on infection | 369 (31.8) | 70 (40.0) | <0.05 |
| Bacterial etiology | 571 (49.1) | 77 (48.4) | 0.933 |
| Viral etiology | 379 (32.6) | 37 (23.3) | <0.05 |
| CNS source | 6 (0.5) | 0 (0.0) | 0.780 |
| ENT source | 268 (23.1) | 18 (11.3) | <0.05 |
| LRTI source | 388 (33.4) | 62 (39.0) | 0.191 |
| Abdominal source | 61 (5.2) | 5 (3.1) | 0.343 |
| GI source | 57 (4.9) | 9 (5.7) | 0.829 |
| UTI source | 203 (17.5) | 15 (9.4) | <0.05 |
| SSTI source | 54 (4.6) | 6 (3.8) | 0.769 |
| Bone or joint source | 9 (0.8) | 0 (0.0) | 0.549 |
| Other source | 57 (4.9) | 5 (3.1) | 0.433 |
| **Lab** |  |  |  |
| Lymphocytes (10<sup>9</sup>/L) | 9.0 [3.1, 18.9] | 6.5 [2.6, 12.7] | 0.389 |
| Creatinine (μmol/L) | 87 [67, 123] | 105 [67, 155] | <0.05 |
| ASAT (U/L) | 28 [21, 45] | 41 [26, 77] | <0.05 |
| ALAT (U/L) | 23 [16, 37] | 29 [18, 52] | <0.05 |
| CRP (mg/L) | 49 [10, 129] | 50 [10, 173] | 0.362 |
| PaO<sub>2</sub> (kPa) | 8.9 [7.2, 11.1] | 11.2 [8.5, 19.0] | <0.05 |
| Lactate (mmol/L) | 1.4 [1.0, 2.2] | 2.5 [1.4, 4.5] | <0.05 |
| **Outcome** |  |  |  |
| Hospitalized | 900 (77.5) | 154 (96.9) | <0.05 |
| ICU within 48h | 0 | 133 (83.6) | <0.05 |
| Deceased within 48h | 0 | 39 (24.5) | <0.05 |

Abbreviations: HEMS: Helicopter Emergency Medical Services, SBP: Systolic Blood Pressure, DBP: Diastolic Blood Pressure, MAP: Mean Arterial Pressure, SpO<sub>2</sub>: Peripheral Oxygen Saturation, GCS: Glasgow Coma Scale, CNS: Central Nervous System, ENT: Ear, Nose, Throat, LRTI: Lower Respiratory Tract Infection, GI: Gastro-intestinal, UTI: Urinary Tract Infection, SSTI: Skin and Soft Tissue Infection, ASAT: Aspartate transaminase, ALAT: Alanine transaminase, CRP: C-Reactive Protein, PaO<sub>2</sub>: Partial Arterial Oxygen Pressure.

### Performance of traditional early warning scores

We first evaluated the performance of two commonly used clinical risk scores, qSOFA and NEWS. qSOFA achieved the highest specificity (0.882, 95% CI: 0.837-0.920), but showed low sensitivity (0.357, 95% CI: 0.174-0.550), which would miss identifying a substantial number of patients who deteriorate (*Table 2*). In contrast, NEWS exhibited high sensitivity (recall 0.786, 95% CI: 0.621-0.927) but at the cost of low specificity (0.506, 95% CI: 0.444-0.567), consistent with its role as a highly sensitive screening tool to predict deterioration. In terms of detection ability, both qSOFA and NEWS demonstrated moderate performance, with AUROC values of 0.693 (95% CI: 0.593-0.786) and 0.712 (95% CI: 0.613-0.799), respectively (*Figure 2A*).

**Table 2:** Performance metrics for qSOFA and NEWS to predict deterioration, defined as ICU admission and/or death within 48 hours after ED admission. The table reports AUROC, actual sensitivity, specificity, PPV, NPV, F1, and F2 scores for each model. Confidence intervals are provided in brackets. qSOFA and NEWS are rule-based clinical scoring systems evaluated at their standard predefined cutoffs (qSOFA ≥ 2; NEWS ≥ 5).

| Model | AUROC | Sensitivity | Specificity | PPV | NPV | F1 | F2 |
| --- | --- | --- | --- | --- | --- | --- | --- |
| qSOFA | 0.693<br>(0.593-0.786) | 0.357<br>(0.174-0.550) | 0.882<br>(0.837-0.920) | 0.263<br>(0.122-0.413) | 0.921<br>(0.883-0.953) | 0.303<br>(0.147-0.447) | 0.333<br>(0.163-0.496) |
| NEWS | 0.712<br>(0.613-0.799) | 0.786<br>(0.621-0.927) | 0.506<br>(0.444-0.567) | 0.158<br>(0.101-0.224) | 0.952<br>(0.911-0.985) | 0.263<br>(0.176-0.354) | 0.438<br>(0.312-0.547) |

Abbreviations: AUROC: Area Under the Receiver Operating Characteristic, PPV: Positive Predictive Value, NPV: Negative Predictive Value, qSOFA: quick Sequential Organ Failure Assessment, NEWS: National Early Warning Score.

**Figure 2:**
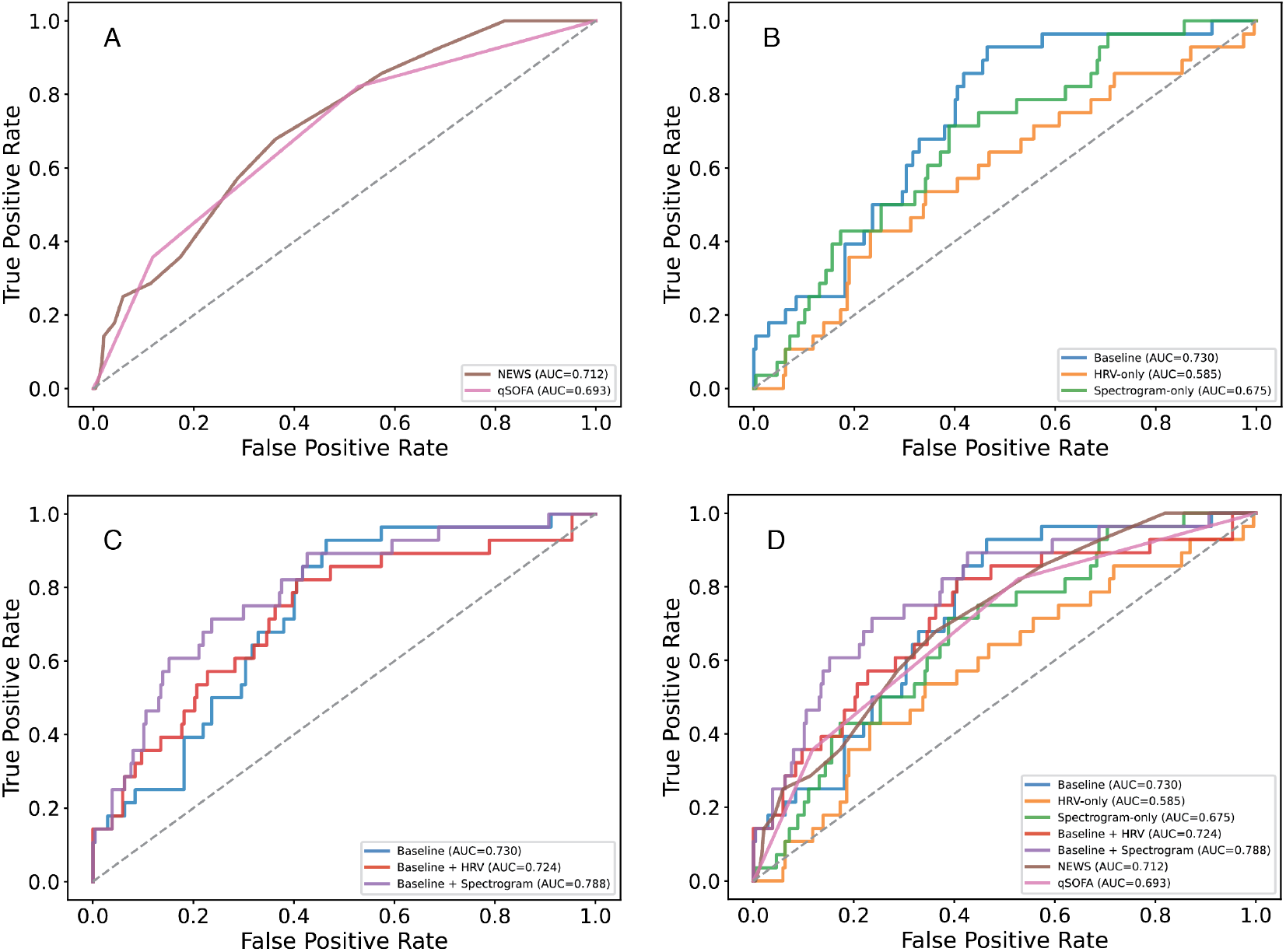
Overview of the Receiver Operating Characteristic (ROC) of the scores and models in the study, including Area Under the ROC (AUROC) for predicting deterioration (ICU admission and/or death) within 48 hours after emergency department admission. A: ROC of the two common early warning scores: National Early Warning Score (NEWS) and quick Sequential Organ Failure Assessment (qSOFA). B: ROC curve of the developed baseline model (age, sex, vitals), solely Heart Rate Variability (HRV) model, and solely Spectrogram model. C: ROC curve of the baseline combined with HRV and baseline combined with Spectrograms multimodal models compared to the baseline model. D: Overview NEWS and qSOFA compared to all models presented in the study.

### Baseline model: age, sex and vital parameters

Next, we developed a baseline model that includes age, sex, and vital signs obtained upon triage. The model trained on baseline data outperformed both the qSOFA and NEWS scoring systems across several metrics, though not uniformly. The baseline model achieved an AUROC of 0.730 (95% CI: 0.640-0.808), indicating better discrimination and better sensitivity-specificity trade-offs than qSOFA and NEWS (*Figure 2B*). At a fixed sensitivity of approximately 0.80, the model achieved a specificity of 0.595 (95% CI: 0.533-0.659). Although precision remained low (0.193, 95% CI: 0.126-0.267), the model showed improved negative predictive value (NPV: 0.966, 95% CI: 0.930-0.993), resulting in comparable F1 scores and higher F2 scores relative to traditional methods (*Table 3*).

**Table 3:** Model performance metrics for the baseline model (age, sex, vitals), HRV, Spectrograms, baseline + HRV, and baseline + Spectrograms to predict deterioration, defined as ICU admission and/or death within 48 hours after ED admission. The table reports AUROC, actual sensitivity, specificity, PPV, NPV, F1, and F2 scores for each model. The highest value in each column is highlighted in bold, and confidence intervals are provided in brackets. The classification threshold was determined on the test set to achieve a sensitivity at or above approximately 0.8. Performance was reported at the ROC operating point closest to, but not below, a target sensitivity of 0.8.

| Model | AUROC | Sensitivity | Specificity | PPV | NPV | F1 | F2 |
| --- | --- | --- | --- | --- | --- | --- | --- |
| Baseline | 0.730<br>(0.640-0.808) | 0.821<br>(0.667-0.955) | 0.595<br>(0.533-0.659) | 0.193<br>(0.126-0.267) | 0.966<br>(0.930-0.993) | 0.313<br>(0.215-0.408) | 0.498<br>(0.373-0.604) |
| HRV-only | 0.585<br>(0.468-0.696) | 0.821<br>(0.667-0.958) | 0.291<br>(0.232-0.350) | 0.120<br>(0.076-0.169) | 0.932<br>(0.872-0.986) | 0.210<br>(0.138-0.284) | 0.380<br>(0.269-0.476) |
| Spectrogram-only | 0.675<br>(0.568-0.768) | 0.821<br>(0.656-0.957) | 0.380<br>(0.319-0.443) | 0.135<br>(0.085-0.189) | 0.947<br>(0.898-0.989) | 0.232<br>(0.154-0.310) | 0.408<br>(0.290-0.507) |
| Baseline + HRV | 0.724<br>(0.613-0.820) | 0.821<br>(0.667-0.957) | 0.595<br>(0.531-0.658) | 0.193<br>(0.127-0.270) | 0.966<br>(0.931-0.993) | 0.313<br>(0.217-0.411) | 0.498<br>(0.371-0.605) |
| Baseline + Spectrogram | **0.788**<br>**(0.690-0.869)** | 0.821<br>(0.667-0.958) | **0.624**<br>**(0.564-0.686)** | **0.205**<br>**(0.133-0.287)** | **0.967**<br>**(0.937-0.993)** | **0.329**<br>**(0.227-0.431)** | **0.513**<br>**(0.385-0.630)** |

Abbreviations: AUROC: Area Under the Receiver Operating Characteristic, PPV: Positive Predictive Value, NPV: Negative Predictive Value, HRV: Heart Rate Variability.

### ECG-based models: HRV-features and spectrograms

We developed two models to evaluate the accuracy of ECG-based models to predict deterioration, being i) a model using ECG-based HRV-features, and ii) a model using ECG-based spectrograms. First, the model based solely on HRV features (HRV-only) exhibited limited predictive power, with an AUROC of 0.585 (95% CI: 0.468-0.696), as shown in *Figure 2B* and *Table 3*. All performance metrics were lower than those of the baseline model, suggesting that the limited performance is likely due to the HRV extraction approach rather than the ECG signal itself, as models based on ECG spectrograms demonstrated improved performance. Next, we developed a model based on ECG spectrograms. The model trained solely on ECG spectrograms achieved better performance than the HRV-only model. The spectrogram-only model achieved an AUROC of 0.675 (95% CI: 0.568-0.768) and a specificity of 0.380 (95% CI: 0.319-0.443) at a fixed sensitivity of 0.8 (*Table 3* and *Figure 2B*). These findings suggest that spectrogram-derived features alone provide limited discriminative performance.

### Multimodal integration of baseline model with ECG-based models

We next combined the baseline model (i.e., age, sex, vital parameters) with the ECG-based models to investigate whether multimodal integration improves predictive performance. First, combining the baseline model with HRV features did not lead to improved specificity (0.595, 95% CI: 0.531-0.658) and F2 score (0.498, 95% CI: 0.371-0.605) compared to the baseline model without HRV features, and minimal changes in PPV and NPV. The AUROC is slightly lower than that of the baseline model (*Figure 2C)*. Next, we integrated the baseline model with ECG spectrograms. This consistently yielded the best overall performance across all calculated metrics. The multimodal “baseline + spectrogram” model achieved the highest AUROC (0.788, 95% CI: 0.690-0.869), and improved specificity (0.624, 95% CI: 0.564-0.686) at fixed sensitivity. This improvement resulted in the highest PPV (0.205, 95% CI: 0.122-0.287), NPV (0.967, 95% CI: 0.937-0.993), F1 (0.329, 95% CI: 0.227-0.431), and F2 (0.513, 95% CI: 0.385-0.630) among all models, indicating an improved balance between sensitivity and specificity. An overview of the ROC curves for qSOFA, NEWS, and all presented models is shown in *Figure 2D*.

### Pairwise bootstrap analysis

Pairwise bootstrap analysis (5,000 resamples) demonstrated that the “Baseline + Spectrogram” model showed significantly higher AUROC than the HRV-only model (∆AUROC = 0.203, 95% CI: 0.091–0.315, p <0.001) and the Spectrogram-only model (∆AUROC = 0.113, 95% CI: 0.009–0.218, p = 0.034), as shown in *Table 4*. Although improvements over the Baseline model (∆AUROC = 0.057, p = 0.072) and the Baseline + HRV model (delta AUROC = 0.064, p = 0.096) did not reach statistical significance, both comparisons showed consistent performance gains.

**Table 4:**
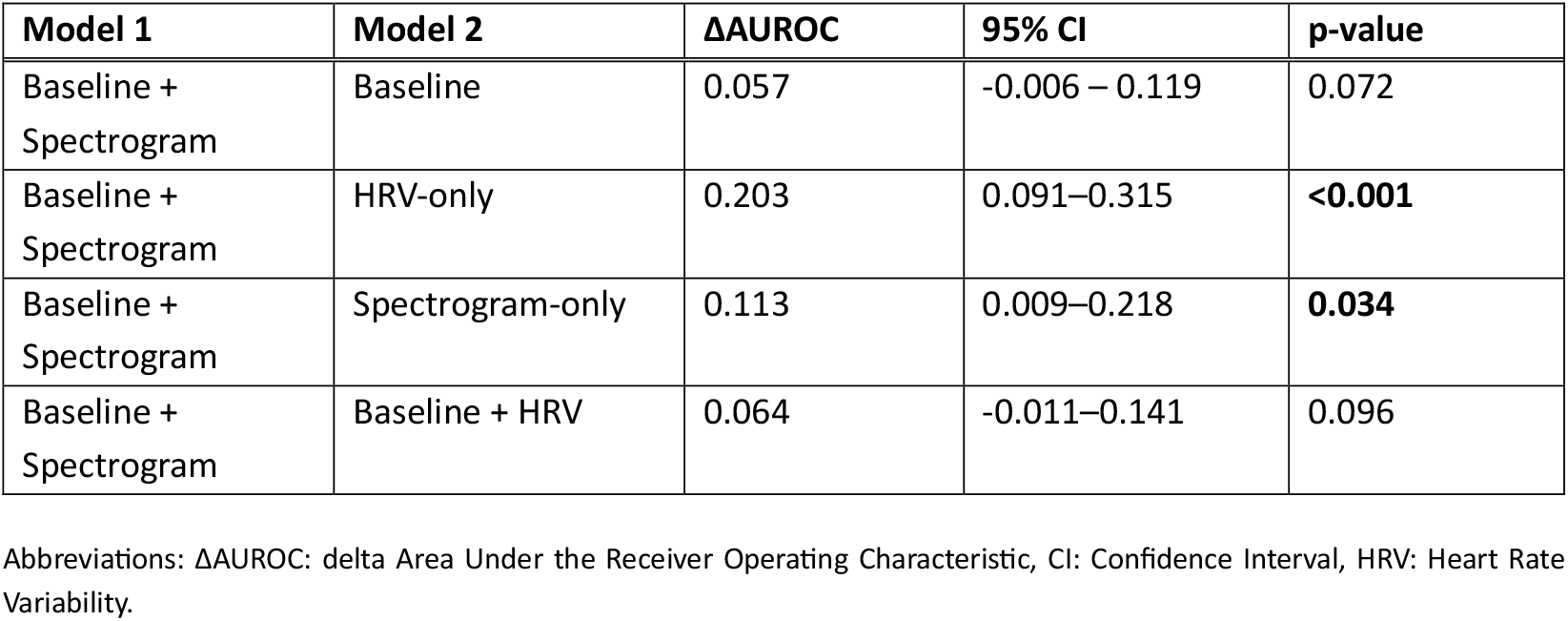
Pairwise bootstrapping, comparing the performance of the “baseline + spectrogram” model to the other presented models: baseline (age, sex, vitals), ECG-derived HRV features (HRV-only), ECG-derived spectrograms (Spectrogram-only), and baseline with HRV combined.

## Discussion

In this study, we compared the predictive performance of two different approaches of using ECG data from the first 20 minutes at the ED compared to current clinical scoring systems in patients with a suspected infection. All models used the same endpoint: clinical deterioration that we defined as ICU admission and/or death within 48 hours of ED admission. The inclusion of a real-world, heterogeneous ED early sepsis patient population reduces the risk of selection bias and enhances generalizability. Our newly developed models were based on either baseline data (i.e., age, sex, vital parameters), HRV features, or complete ECG spectrograms, and we developed multimodal models that combined these models. The baseline model performed better than qSOFA and NEWS scores, showing that nonlinear patterns in routine triage data add predictive value. The ECG spectrogram-only model performed better than the HRV-only model, suggesting that time-frequency ECG information is more informative than HRV features alone. The best performance for predicting clinical deterioration came from the multimodal baseline combined with the ECG spectrogram model, which achieved the highest AUROC, specificity, and F-scores. Together, our results show that ECG spectrogram features derived from continuous early ED ECG-recordings provide complementary predictive information beyond traditional triage data and HRV analysis, improving both discriminative performance and sensitivity-specificity balance compared to vital sign-based models and established clinical scores. The observation that spectrograms are better predictors for deterioration than HRV features suggests that relevant information is lost in processing ECGs to acquire HRV features. Spectrograms retain richer signal characteristics than discrete HRV features, as they encode time-frequency dynamics without predefined feature extraction. Although it is currently unknown whether shorter or longer segments would improve spectrogram-based models, in raw waveform-based arrhythmia classification, an optimum was found at 10-20 minutes of ECG signal, which is comparable to the 20-minute segments in our study [28]. Our study suggests that current techniques do not fully exploit the potential of ECG recordings during the initial period after admission to the ED. This was also shown in a study by Lin et al., which demonstrated that a deep-learning-based ECG processing pipeline performs better at predicting 1-year mortality than models using manually extracted features [29]. Although the lack of explainability potentially hampers adoption by end-users, despite the good performance of the model, it does show the potential of spectrograms and the extra layer of information in it.

Spectrogram analysis of continuous ECG recordings is an emerging method for detecting abnormalities and diseases, and it has been studied especially in the field of arrhythmia detection [23,24]. Using spectrograms proved valuable for reducing raw signals into a single multi-dimensional representation while preserving relevant information. No other studies developed models that classify spectrograms to predict deterioration of early sepsis patients at the ED. The spectrogram generation, leveraging the Short-time Fourier Transform, is, however, methodologically comparable to other ECG spectrogram studies, thereby demonstrating the technical validity and reproducibility of our signal processing approach [23,24,30]. These studies often used 10 to 60 seconds of ECG recordings, while our study used 20 minutes, which allows for capturing slower processes and deteriorating trends. When comparing the spectrogram-based method to other methods to predict deterioration in patients with early sepsis, such as NEWS, qSOFA, and HRV, this study presented models with AUCs in the same range as presented here (0.65-0.89) [17,31–33].

The previously published TREWScore [34], designed to identify patients at risk of developing septic shock, has an endpoint comparable to that of our study. It achieved an AUC of 0.83, with a sensitivity of 0.85 and a specificity of 0.6, slightly higher than the performance metrics of our “Baseline + Spectrogram” model. However, the TREWScore relies on a broad set of features extracted from MIMIC-II [35,36], an ICU-focused public dataset, and incorporates not only vital signs but also laboratory and clinical parameters. Our study demonstrates that comparable performance can be achieved using only baseline characteristics (age, sex, triage vitals) and ECG spectrograms. Furthermore, we aimed to extend these baseline parameters with ECG-derived features to demonstrate improved performance.

A study by Chang et al [38] compared a CNN-based 12-lead ECG model with NEWS for predicting 3-day mortality in emergency department patients, an endpoint similar to ours. They found that the ECG model achieved an AUC similar to NEWS (both 0.85), that combining the ECG model with NEWS reduced performance (AUC 0.83), and that integrating the ECG model with multiple early warning scores (EWS) scores improved AUC (AUC 0.92). These findings highlight the added value of incorporating ECG data alongside routinely collected vital signs at baseline, consistent with the results of our study. The potential of multimodal models combining ECG waveform data with tabular data, as presented in this study, has been shown before. In a recent study on predicting the outcome of ICU patients, the multimodal fusion model showed performance improvement over only tabular data, and it outperformed clinicians and large language models [39]. From a technical perspective, the model we present is different, but it shows the potential of such models and the added value of adding ECG waveforms to tabular data, similar to what we have shown in this study.

### Clinical relevance

This study shows that ECG spectrogram models perform better than the current clinically used EWS. Furthermore, they provide clinically meaningful information beyond traditional vital signs and HRV analysis [17], capturing the full frequency spectrum of cardiac electrical activity in patients with early sepsis at risk of deterioration. As ECG signals are routinely collected and increasingly available through continuous bedside monitoring and wearable devices, spectrogram-based analysis is highly scalable and clinically feasible because it relies solely on bedside monitor data, unlike EWS, which often requires manually collected data. Such models could subsequently serve as clinical decision support tools, assisting physicians in making informed decisions about treatment and discharge following ED admission.

### Limitations

Several limitations should be acknowledged. First, because this analysis uses a CNN to classify spectrograms, it is unclear which specific ECG features contributed to the model’s decisions regarding deterioration. When applying this method in a clinical setting, a deeper understanding of the (patho)physiological elements is preferred. In future studies, this could be partially achieved with explainable AI [40]. Further, we did not determine whether the model used elements related to physiological changes or external effects, such as technical artefacts, that contribute to differences in the ECG spectrogram. To limit potential bias due to technical differences (e.g, different devices, rooms, background noise, etc.), data was collected in a wide variety of patients with suspicion of infection, which is a representative population of patients arriving at the ED with a suspicion of infection. Finally, due to its single-center design, the study lacks external validation. Although we present a proof of concept, future studies should include external validation.

### Future implications

Future studies could focus on exploring the contributing features in spectrograms that provide additional information compared with EWS, triage vitals, and HRV and extend this with raw-waveform-based models. Studies could focus on model performance using spectrograms compared to models using raw waveforms as their input. At the same time, future models that predict patient outcome at the ED could involve spectrograms as their predictive ability showed improved performance over vitals and HRV features. In that context, spectrograms derived from continuous ECG recordings could be deployed at the ED to support physicians in their clinical decision-making. The technique is not yet ready for clinical implementation; external and clinical validation are needed, and a real-time waveforms processing infrastructure needs to be available in the clinic, using data from electronic health records and bedside patient monitors [41]. Furthermore, a better understanding of the characteristics of ECGs that are key to predicting deterioration could help convince clinicians to adopt a spectrogram-based prediction model. When specific characteristics can be identified, the need for full spectrogram analysis in the ED may be lower, for example, if relevant characteristics can be extracted using conventional signal analysis techniques.

### Conclusion

This study demonstrates that ECG spectrograms derived from early ED recordings combined with baseline triage parameters improve early risk stratification for clinical deterioration compared with standard early warning scores and HRV-based models, in patients presenting with suspected infection at the ED. Multi-modal models incorporating spectrograms outperform those based solely on vital signs, EWS, and HRV features. Although current limitations regarding interpretability and validation remain, spectrogram-based analysis represents a promising approach for enhancing early risk stratification and supporting clinical decision-making for patients with suspicion of infection in the ED.

## Data Availability

The data from the Acutelines data- and biobank are not publicly available, but can be accessed upon reasonable request.

## Acknowledgements

The data used in this manuscript are provided by Acutelines. The authors would like to thank Acutelines and all its participants. The establishment of Acutelines has been made possible by funds from the University Medical Center Groningen.

## Statements & Declarations

### Funding

The authors declare that no relevant funds, grants, or other support were received for the preparation of this manuscript. The Acutelines data- and biobank has been made possible by funds from University Medical Center Groningen.

### Competing Interests

The authors have no relevant financial or non-financial interests to disclose.

### Author Contributions

**RJvW:** Conceptualization, Data curation, Investigation, Methodology, Writing – original draft, review, and editing. **ADS:** Investigation, Validation, Writing – review and editing. **LdV:** Conceptualization, Formal Analysis, Investigation, Methodology, Software, Writing – review and editing. **STH:** Investigation, Validation, Writing – review and editing. **CG:** Investigation, Validation, Writing – review and editing. **JMLA:** Methodology, Validation, Writing – review and editing. **NS:** Methodology, Validation, Writing –review and editing. **JCtM:** Conceptualization, Funding acquisition, Resources, Supervision, Writing –review and editing. **HRB:** Conceptualization, Funding acquisition, Resources, Supervision, Writing –review and editing. **JL:** Formal Analysis, Methodology, Software, Visualization, Writing – original draft, review, and editing.

### Ethics Approval

This study and the Acutelines data- and biobank were reviewed and approved by the institutional research board of the University Medical Center Groningen (#18939). The Acutelines data- and biobank is registered under NCT04615065 at ClinicalTrials.gov.

### Consent

Participants were asked for written informed consent, when applicable by proxy or opt-out.

## Notes

### Competing Interest Statement

The authors have declared no competing interest.

### Author Declarations

Ethics Committee of University Medical Center Groningen gave ethical approval for this work

