## Supplementary material for "ECG spectrogram-based deep learning model to predict deterioration of patients with early sepsis at the emergency department: a study from the Acutelines data- and biobank"

### Supplemental material

#### A. Detailed methods

*ECG spectrogram encoder*

To capture time-frequency characteristics of ECG signals, raw ECG waveforms were transformed into spectrogram representations. Spectrograms were generated with a Tukey window, a step size of 5 seconds, 50% overlap, and were log-transformed. These spectrograms were processed using a convolutional neural network (CNN)-based encoder, which is particularly suited for learning local patterns and hierarchical representations in two-dimensional structured data.

The CNN encoder comprises four sequential convolutional blocks (32–64–128–256). Each block contains a 2D convolution layer with a 3x3 kernel and unit padding, batch normalization (BN), and a Sigmoid Linear Unit (SiLU) activation function. Spatial downsampling is conducted using a 2x2 max pooling after the first three blocks, while the final block preserves the spatial resolution to maintain higher-level feature representations. An adaptive average pooling layer is applied to aggregate features across time and frequency dimensions, followed by a fully connected projection layer to produce a fixed-length embedding. This module aims to capture latent physiological dynamics that are not directly observable from summary clinical measurements.

*Vital signs and HRV feature encoder*

Structured physiological variables, including baseline age, sex, and routine vital signs, and 38 ECG-derived HRV features that were calculated using Neurokit 2 [1] (full list in Supplemental table A1), were fed into a multi-layer perceptron (MLP)-based encoder. Given the low dimensionality and non-spatial nature of these features, MLPs provide an efficient and stable modeling choice. The MLP encoder consists of two fully connected layers (input-dim–128–64) with non-linear SiLU activations, with dropout regularization applied after the first hidden layer. For ECG-derived HRV features, missing values were imputed with zeros, and an additional binary mask variable was appended to indicate feature availability. As all features were standardized using training-set statistics, zero corresponds to the population mean and provides a stable, fold-consistent placeholder, while the mask allows the model to explicitly capture the potentially informative nature of HRV missingness. This design enables the model to learn to down-weight or ignore unreliable features rather than excluding patients from analysis.

*Multimodal feature fusion and prediction*

For the multimodal model, embeddings generated by the CNN-based spectrogram encoder and the MLP-based encoder were concatenated at the feature level. The fused embeddings were subsequently processed by a multi-layer fusion head to learn cross-modality interactions, which comprises two fully connected layers with SiLU activations and dropout regularization (input-dim–128–64), followed by a final linear layer yielding a single logit. This fusion head was the same as the single-modality MLP to make the prediction comparable. The output logit was converted to a probability via a sigmoid activation for binary classification.

*Training and optimization*

All models were trained using binary cross-entropy loss with logits, with a positive-class weighting factor to account for class imbalance in the training data. Model parameters were optimized using the AdamW optimizer, with weight decay applied to mitigate overfitting. Gradient norm clipping was employed to stabilize training. Input features were normalized using statistics (mean and standard deviation) computed solely from the training set to avoid information leakage. Models were trained for up to 60 epochs, with early stopping based on the validation F1 score (patience = 8 epochs). The optimal model was selected according to the best validation F1 score, reflecting a balance between sensitivity and precision in the presence of class imbalance. Dropout regularization was applied in training, and batch normalization layers were used to improve optimization stability.

*Model variants and ablation design*

The modular architecture enables systematic ablation experiments under a unified framework. By selectively enabling and disabling individual encoders, single-modality baselines (baseline-only, spectrogram-only, HRV-only, and Vitals-HRV models) were directly compared with the multimodal model. All models share identical optimization strategies, loss functions, and evaluation procedures, ensuring that performance differences are attributable to modality inclusion rather than differences in training configuration.

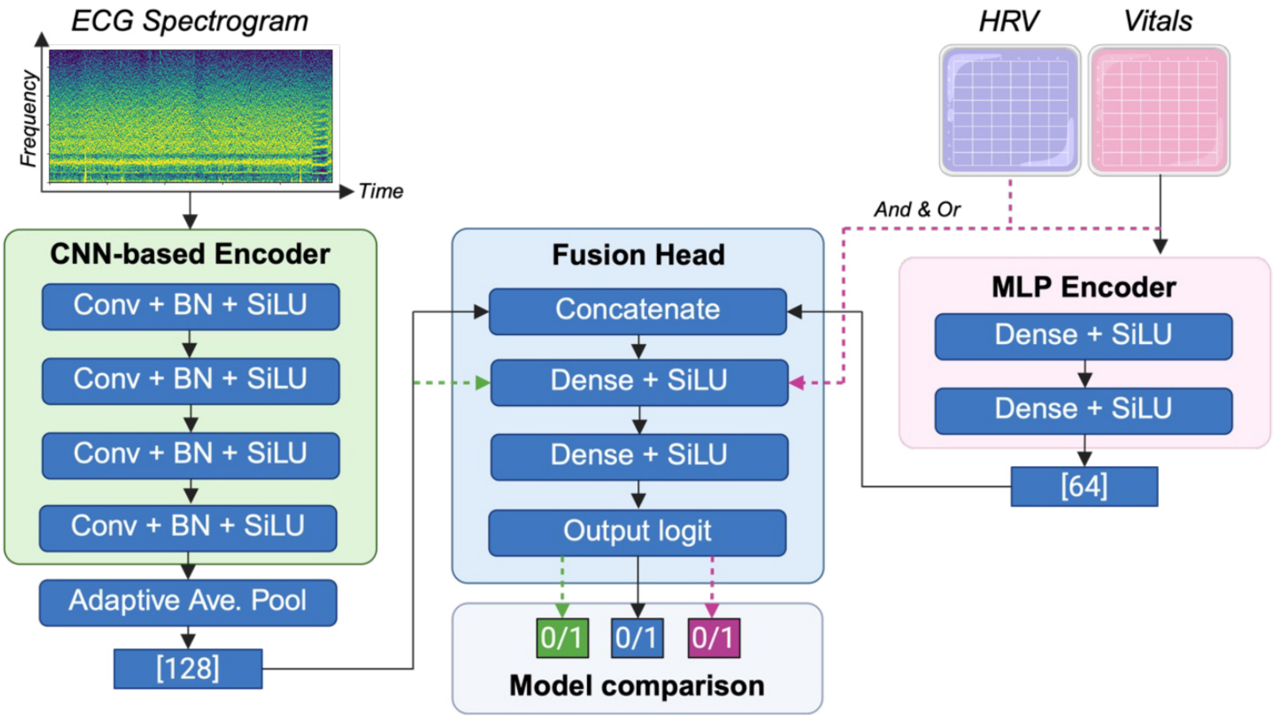


Figure A1: Model architecture, showing the MLP for baseline and HRV data and CNN for spectrogram data. HRV: Heart Rate Variability, CNN: Convolutional Neural Network, Conv: Convolutional Block, BN: batch normalization, SiLU: Sigmoid Linear Unit

Table A1: Overview of HRV features included in the HRV model. Descriptions adapted from Neurokit2 documentation [1,2].

| Name | Domain | Description |
|---|---|---|
| MeanNN | time | Mean of RR intervals (also known as AVNN) |
| SDNN | time | Standard deviation of RR intervals, overall |
| SDANN1 | time | Standard deviation of RR intervals, over 1 minute |
| SDNNI1 | time | Mean of standard deviation of RR intervals, over 1 minute |
| SDANN2 | time | Standard deviation of RR intervals, over 2 minutes |
| SDNNI2 | time | Mean of standard deviation of RR intervals, over 2 minutes |
| SDANN5 | time | Standard deviation of RR intervals, over 5 minutes |
| SDNNI5 | time | Mean of standard deviation of RR intervals, over 5 minutes |
| RMSSD | time | Root mean square of successive differences |
| SDSD | time | Standard deviation of successive differences |
| CVNN | time | Coefficient of variation of RR intervals (SDNN divided by MeanNN) |
| CVSD | time | Coefficient of variation of successive differences (RMSSD divided by MeanNN) |
| MedianNN | time | Median of RR intervals |
| MadNN | time | Median of absolute deviation |
| MCVNN | time | MadNN divided by MeanNN |
| IQRNN | time | Interquartile range |
| SDRMSSD | time | SDNN divided by RMSSD |
| Prc20NN | time | 20 percentile of RR intervals |
| Prc80NN | time | 80 percentile of RR intervals |
| pNN50 | time | Percentage of difference between RR intervals >50ms |
| pNN20 | time | Percentage of difference between RR intervals >20ms |
| MinNN | time | Minimum RR intervals |
| MaxNN | time | Maximum RR intervals |
| HTI | time | HRV triangular index |
| TINN | time | Baseline width of RR interval distribution plot |
| ULF | frequency | Ultra low frequency bandpower (0-0.0033 Hz) |
| VLF | frequency | Very low frequency bandpower (0.0033-0.04 Hz) |
| LF | frequency | Low frequency bandpower (0.04-0.15 Hz) |
| HF | frequency | High frequency bandpower (0.15-0.4 Hz) |
| VHF | frequency | Very high frequency bandpower (0.4-0.5 Hz) |
| TP | frequency | Total power |
| LFHF | frequency | LF divided by HF ratio |
| LFn | frequency | Normalized LF power |
| HFn | frequency | Normalized HF power |
| LnHF | frequency | Log transformed HF power |
| SD1 | non-linear | Standard deviation 1 of Poincaré plot |
| SD2 | non-linear | Standard deviation 2 of Poincaré plot |
| SampEn | non-linear | Sample entropy |

#### B. Tree-based models

In the main analysis for tabular data, we used an MLP model. However, tree-based models are commonly the most widely used for tabular data. To see whether there were performance differences using tree-based models and MLP for tabular data, we applied XGBoost and LightGBM to the baseline (age, sex, vitals), HRV-only, and baseline + HRV scenarios. Most performed equally or slightly worse than their MLP-based equivalent, except for the HRV-only model (AUC 0.613 with XGBoost compared to 0.585 with MLP) and Baseline + HRV (AUC=0.753 with XGBoost compared to 0.724 with MLP), as can be seen in *Table B1* and *Figure B1*. We therefore decided to perform no additional analysis with tree-based models in the main manuscript.

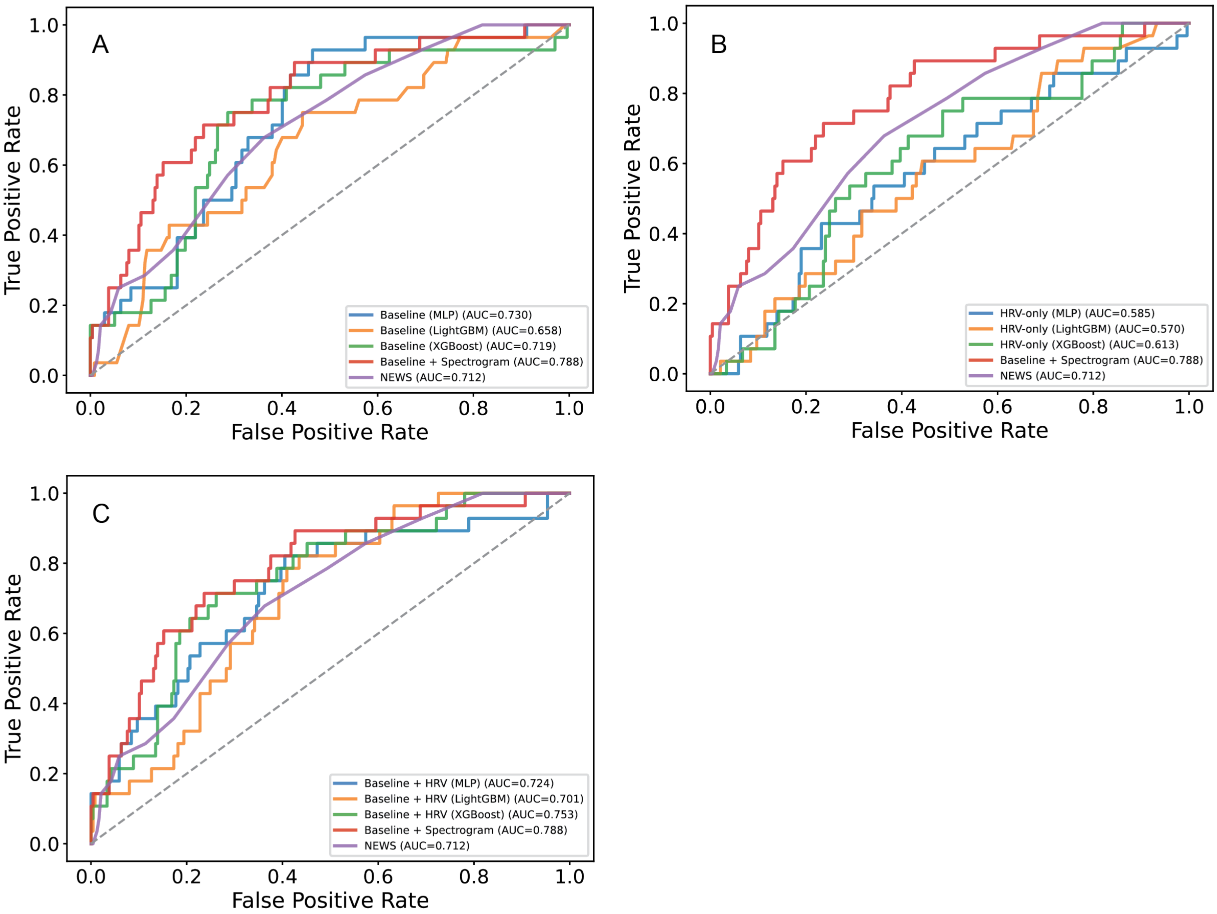


Figure B1: Comparison of receiver operating characteristic (ROC) between tree-based models trained on tabular data, the multimodal "baseline + Spectrogram" model, and the NEWS score.

Table B1: Overview of tree-based metrics for tabular data at the fixed sensitivity of 0.8 on test set. Metrics are reported with confidence intervals between brackets.

| **Model** | **AUROC** | **Sensitivity** | **Specificity** | **PPV** | **NPV** | **F1** | **F2** |
|---|---|---|---|---|---|---|---|
| Baseline (LightGBM) | 0.658 (0.550-0.758) | 0.821 (0.667-0.957) | 0.338 (0.274-0.397) | 0.128 (0.081-0.179) | 0.941 (0.887-0.988) | 0.221 (0.146-0.296) | 0.394 (0.277-0.496) |
| Baseline (XGBoost) | 0.719 (0.608-0.812) | 0.821 (0.667-0.960) | 0.591 (0.527-0.649) | 0.192 (0.124-0.264) | 0.966 (0.931-0.993) | 0.311 (0.210-0.407) | 0.496 (0.367-0.605) |
| HRV-only (LightGBM) | 0.570 (0.462-0.671) | 0.857 (0.706-0.966) | 0.308 (0.248-0.369) | 0.128 (0.082-0.180) | 0.948 (0.892-0.988) | 0.222 (0.147-0.300) | 0.400 (0.284-0.497) |
| HRV-only (XGBoost) | 0.613 (0.508-0.708) | 0.857 (0.714-0.969) | 0.224 (0.168-0.276) | 0.115 (0.075-0.163) | 0.930 (0.857-0.984) | 0.203 (0.136-0.276) | 0.375 (0.269-0.473) |
| Baseline + HRV (LightGBM) | 0.701 (0.615-0.776) | 0.821 (0.667-0.962) | 0.565 (0.500-0.628) | 0.183 (0.118-0.252) | 0.964 (0.929-0.993) | 0.299 (0.204-0.391) | 0.483 (0.352-0.588) |
| Baseline + HRV (XGBoost) | 0.753 (0.657-0.833) | 0.821 (0.667-0.957) | 0.578 (0.515-0.640) | 0.187 (0.122-0.263) | 0.965 (0.931-0.993) | 0.305 (0.208-0.403) | 0.489 (0.361-0.598) |
